# Nasopharyngeal Angiotensin Converting Enzyme 2 (ACE2) Expression as a Risk-Factor for SARS-CoV-2 Transmission in Concurrent Hospital Associated Outbreaks

**DOI:** 10.1101/2023.03.20.23287264

**Authors:** Aidan M. Nikiforuk, Kevin S. Kuchinski, Katy Short, Susan Roman, Mike A. Irvine, Natalie Prystajecky, Agatha N. Jassem, David M. Patrick, Inna Sekirov

**Affiliations:** British Columbia Centre for Disease Control, Vancouver, BC V5Z 4R4, Canada; Department of Pathology and Laboratory Medicine, University of British Columbia, Vancouver, BC V6T 1Z4, Canada; Fraser Health Authority, New Westminster, BC V3L 3C2, Canada; School of Population and Public Health, University of British Columbia, Vancouver, BC V6T 1Z4, Canada; Faculty of Health Sciences, Simon Fraser University, Burnaby, BC V5A 1S6, Canada

**Keywords:** ACE2, SARS-CoV-2, Infection Tracing, Transmission Network, Outbreak Investigation, Multivariable Analysis, Poisson Regression Model, Negative Binomial Regression

## Abstract

**Background:** Widespread human-to-human transmission of the severe acute respiratory syndrome coronavirus two (SARS-CoV-2) stems from a strong affinity for the cellular receptor angiotensin converting enzyme two (ACE2). We investigate the relationship between a patient’s nasopharyngeal *ACE2* transcription and secondary transmission within a series of concurrent hospital associated SARS-CoV-2 outbreaks in British Columbia, Canada.

**Methods:** Epidemiological case data from the outbreak investigations was merged with public health laboratory records and viral lineage calls, from whole genome sequencing, to reconstruct the concurrent outbreaks using infection tracing transmission network analysis. *ACE2* transcription and RNA viral load were measured by quantitative real-time polymerase chain reaction. The transmission network was resolved to calculate the number of potential secondary cases. Bivariate and multivariable analyses using Poisson and Negative Binomial regression models was performed to estimate the association between *ACE2* transcription the number of SARS-CoV-2 secondary cases.

**Results:** The infection tracing transmission network provided n = 76 potential transmission events across n = 103 cases. Bivariate comparisons found that on average *ACE2* transcription did not differ between patients and healthcare workers (P = 0.86). High *ACE2* transcription was observed in 98.6% of transmission events, either the primary or secondary case had above average *ACE2*. Multivariable analysis found that the association between *ACE2* transcription and the number of secondary transmission events differs between patients and healthcare workers. In health care workers Negative Binomial regression estimated that a one unit change in *ACE2* transcription decreases the number of secondary cases (B = −0.132 (95%CI: −0.255 to −0.0181) adjusting for RNA viral load. Conversely, in patients a one unit change in *ACE2* transcription increases the number of secondary cases (B = 0.187 (95% CI: 0.0101 to 0.370) adjusting for RNA viral load. Sensitivity analysis found no significant relationship between *ACE2* and secondary transmission in health care workers and confirmed the positive association among patients.

**Conclusion:** Our study suggests that *ACE2* transcription has a positive association with SARS-CoV-2 secondary transmission in admitted inpatients, but not health care workers in concurrent hospital associated outbreaks, and it should be further investigated as a risk-factor for viral transmission.

## Introduction

The severe acute respiratory syndrome coronavirus 2 (SARS-CoV-2) causes potentially life threatening lower respiratory and systemic inflammatory disease defined as COVID-19[1–3]. SARS-CoV-2 has spread widely since late 2019 causing a global pandemic. Two previous public health emergencies have provided the opportunity to study human-to-human transmission of pathogenic coronaviruses. In 2003, the severe acute respiratory syndrome emerged infecting eight-thousand four-hundred and twenty-two people [4]. In 2012 and 2015 the Middle East respiratory syndrome coronavirus (MERS-CoV) caused epidemics in the Kingdom of Saudi Arabia and South Korea with two-thousand five hundred and sixty-two laboratory confirmed cases reported to the World Health Organization[5]. SARS- and MERS-CoV spread predominately in health care settings and community spread was controlled by public health interventions or self-limited[4,5]. The transmission pattern of SARS-CoV-2 contrasts that of the other pathogenic coronaviruses (SARS-CoV and MERS-CoV). SARS-CoV-2 has predominately spread within the community, the virus has evolved to become more infectious over time and vaccination does not protect against re-infection or transmission of newer strains [6].

Transmission of respiratory viruses like SARS-CoV-2 involves a complex interplay of social, environmental, and biological variables[7]. Numerous observational studies have described risk factors of SARS-CoV-2 transmission across a variety of settings including hospitals[8], households[9], and schools[10,11]. Cumulative evidence suggests that the increased human-to-human transmissibility of SARS-CoV-2 in comparison to SARS-CoV stems from stronger affinity for the primary host receptor angiotensin converting enzyme 2 (ACE2)[12–14]. SARS-CoV-2 binds to ACE2 through its spike glycoprotein, the spike protein undergoes proteolytic cleavage before binding with ACE2 prompting endocytosis and membrane fusion[13]. The Omicron variant of SARS-CoV-2 has a stronger affinity for ACE2 and less reliance on host proteases biasing cell entry towards endocytosis, consequently less cell fusion and syncytia formation occurs[15]. ACE2 is highly expressed in the upper and lower respiratory tract, upregulation may occur in response to stimulation with interferon[16]. SARS-CoV-2 exploits upregulation of *ACE2* in an interferon rich environment (like the early stages of an innate immune response) to spread from cell-to-cell producing a high viral titre[17]. In the later stages of replication, SARS-CoV-2 downregulates *ACE2*, preventing any benefits to the host mediated by interferon stimulated upregulation[18,19]. SARS-CoV-2 has a shifting relationship with ACE2 and like viral load, ACE2 expression fluctuates through the course of viral infection[20– 22]. Regardless, SARS-CoV-2 replicates most efficiently in tissues rich with ACE2, the strong positive association between viral load and ACE2 expression at the time of diagnostic testing increase the degree of viral shedding and thereby, the risk of ongoing transmission [23].

To understand the role of *ACE2* expression in SARS-CoV-2 transmission, we conducted an infection tracing network analysis of positive testers in concurrent, single-site, hospital associated COVID-19 outbreaks in British Columbia during late 2020 and early 2021. The study aims to: (i) describe the outbreaks in context of person, place, and time, (ii) visualize an infection tracing transmission network to infer the number of secondary cases per primary case and (iii) quantify the relationship between the number of potential secondary cases (outcome) and nasopharyngeal transcription of transmembrane ACE2 (exposure) while adjusting for viral RNA load, and interaction by case designation (patient or healthcare worker). We build from previous work which demonstrated no relationship between transmembrane nasopharyngeal *ACE2* transcription and age, biological sex or *TMPRSS2* transcription in a sample of COVID-19 negative persons[23]. This study contributes novel evidence of how nasopharyngeal *ACE2* transcription may drive SARS-CoV-2 transmission and highlights the potential role of respiratory masks and infection prevention and control measures in limiting the viruses’ nosocomial spread.

## Methods

### Study Design and Participants

We conducted surveillance for COVID-19 cases in a British Columbian hospital over a series of outbreaks, from declaration of the first facility associated outbreak on 07-11-2020 to the date the last outbreak was declared over on 04-01-2021 (dd-mm-yyyy). Participants included patients and health care workers who tested positive for COVID-19 upon self-reporting or showing indicative symptoms (e.g. fever, fatigue, cough and loss of taste or smell) or through asymptomatic point prevalence testing during the outbreak investigations. Inclusion criteria were applied to select study participants whose diagnostic specimens were tested centrally at the British Columbia Center for Disease Control Public Health Laboratory, underwent SARS-CoV-2 whole genome sequencing, and had adequate remaining volume of nucleic acid extract to assay *ACE2* transcription. In the case that a participant was tested for COVID-19 more than once, their first diagnostic specimen from the study period was sampled. Participants who met the inclusion criteria (n = 202) were excluded from the study if their specimen collection container was not identifiable (n = 22), whole genome sequencing failed and was not able to classify the viral lineage (n = 63) or *ACE2* transcription was unmeasurable (n = 14) (Figure S1). An analytic dataset of n= 103 participants was used for analysis. Demographic variables of age, biological sex, case description (patient or health care worker) and site of infection/site of transmission (hospital unit) were drawn from public health laboratory data or the outbreak report of the investigating epidemiologist. Outbreaks were defined by the presence of one or more confirmed cases of COVID-19 which were epidemiologically linked within hospital units (Table S3). The laboratory procedures were performed in a laboratory accredited by the College of American Pathologists and BC’s Diagnostic Accreditation Program using validated RT-qPCR and whole genome sequencing protocols[24–26].

### Ethics Statement

Ethical approval for the study was sought from the University of British Columbia human ethics board (#H20-01110) which was harmonized with the Fraser Health Authority. Written informed consent was not required. All data was de-identified prior to analysis and the results were not linked back to any identifying records. This study was deemed as minimal risk to the participants involved. To ensure privacy the site of the outbreak series will not be disclosed.

### Procedures

Participant’s diagnostic specimens were collected by nasopharyngeal swab and stored in Universal Transport Medium™ (UTM®) (COPAN) and stored at 4LC before RNA extraction. RNA extraction was performed on the MagMAX-96™ platform with the Viral RNA isolation kit (ThermoFisher). Host and viral gene transcription was assayed by quantitative real-time polymerase chain reaction (RT-qPCR) on the Applied Biosystems 7500 Real-Time PCR platform using TaqMan FastVirus 1-step polymerase (ThermoFisher). Reaction volumes were 20 μl, with 5 μl of RNA template, 1μl of 20X primer/probe, 5 μl FastVirus and 9 μl of nuclease free water. Cycling conditions were set to: 50□C for 5min, 95□C for 20s followed by 40 cycles of 95□C for 15s and 60□C for 1 min. As previously described, multiplex RT-qPCR reactions were used to detect host (*ACE2, GAPDH, RNaseP*) and viral (*E* gene) transcription[27]. Participants were diagnosed COVID-19 positive with an *E* gene cycle threshold value < 38 and a *RNaseP* gene cycle threshold value < 40. The *E* gene Ct values were transformed to genome equivalents per millilitre using a standard curve of SARS-CoV-2 synthetic RNA (MN908947.3) (Twist Bioscience)[23]. Relative gene transcription for *ACE2* was calculated in proportion to the control gene *GAPDH* using the 2^−^ΔΔ^Ct^ method[28]. SARS-CoV-2 whole genome sequencing was performed on all diagnostic specimens which tested positive for SARS-CoV-2 by RT-qPCR. Viral genomes were amplified using a 1200bp amplicon scheme and sequenced on an Illumina NextSeq instrument[24]. Genome assembly was performed via a modified ARTIC Nextflow pipeline[29]. Quality control did not pass any genomes with < 85% completeness or < 10X depth of coverage. Viral lineages were assigned using the PANGOLIN tool (Version 1.15.1)[30]. All molecular and genomic testing was performed at the BCCDC-PHL.

### Variable Definition

Demographic variables of interest were drawn from public health laboratory data, or the outbreak reports provided by the investigating epidemiologist. The measures included in the study are age, biological sex, case description, diagnostic specimen collection date (dd-mm-yyyy), SARS-CoV-2 *E* gene cycle-threshold (Ct) value, *ACE2* gene transcription, SARS-CoV-2 lineage (PANGO lineage), site of transmission (hospital unit) and site of infection (hospital unit). Age, *E* gene Ct value (transformed to log10 GE/mL[23]) and *ACE2* gene transcription are continuous numeric variables. Biological sex, case description, viral lineage, collection date and site of transmission/infections are categorical variables. We assumed that collection date equals date of symptom onset for this study because once the hospital declared an outbreak, patients and health care workers were required to be tested at the onset of symptoms. *ACE2* gene transcription was transformed to a categorical value using the mean transcription value (XL = 0.00, SD = 1.08) of COVID-19 negative testers, from a previous study which collected a random sample of n = 212 nasopharyngeal specimens in British Columbia during 2020[23]. The number of potential secondary cases per primary case was determined by infection tracing transmission network analysis. *ACE2* gene transcription and number of potential secondary cases are the exposure and outcome of interest. Viral load and case description were included in multivariable analysis because, they meet the definition of a confounder and are a common cause of the exposure (*ACE2* gene transcription) and outcome (number of secondary cases)[31]. Viral load and *ACE2* gene transcription share a strong positive association, people with high *ACE2* gene transcription were found to have high viral loads[32]. Viral load relates to transmission in that people with high viral loads may shed more infectious virus[33,34]. Case description (patient or healthcare worker) relates to *ACE2* gene transcription because patients may have comorbidities or receive medications which affect *ACE2* transcription[35]. Health care workers have additional social connections relative to in-patients, as they can leave work and be exposed to SARS-CoV-2 outside of clinical areas or in the community[8].

### Statistical Analysis

#### Descriptive Statistics

The analytic data (n = 103) was used for bivariate analysis. The variables of interest were stratified by case description and categorical *ACE2* transcription. An epidemiological curve was visualized for the series of concurrent hospital outbreaks from 07-11-2020 to 04-01-2021 (dd-mm-yyyy). Parametric statistical tests were used given the large sample size of our study[36].

#### Infection Tracing Network Analysis

A matrix of all possible transmission pairs was permuted using SARS-CoV-2 lineage classifications, for two cases to form a pair they had to share the same lineage. SARS-CoV-2 lineages circulating in the hospital were diverse enough that the minimal difference of single nucleotide polymorphisms between them was unlikely to have occurred by mutation during a single transmission event, suggestion multiple independent introductions (Table S4). Four assumptions (i-iv) were used to select potential transmission pairs from all possible permutations. The first assumption (i) stipulated that specimen collection date (symptom onset date) of the primary case was before that of the secondary case. The second assumption (ii) affirms that the primary case’s unit of transmission equals the secondary case’s unit of infection. The third assumption (ii) holds that the collection date of the secondary case is at least one-serial interval (5 days) from that of the primary case[37]. The fourth assumption (iv) dictates that the collection date of the secondary case is not more than three-serial intervals (15 days) from the primary case[38]. Potential transmission pairs, which met the four-assumptions, were stratified by categorical *ACE2* transcription (High/Low) or viral lineage and plotted overtime.

#### Primary Analysis

Multivariable analysis was performed to estimate the relationship between *ACE2* transcription and number of secondary cases using a Poisson generalized linear regression model. Variable importance was assessed conceptually using the common cause criterion and statistically by the partial F-test. Collinearity was measured using the variable inflation factor, variables with a value greater than five were excluded[39]. Effect modification terms were included if they were statistically significant and supported conceptually. The two assumptions of Poisson models; overdispersion and zero inflation were checked using a ratio of residual deviance to degrees of freedom (theta)[40] and the score test[41], respectively. If the specified Poisson model failed to meet either of these assumptions than an alternative quasi-Poisson or Negative Binomial (NB) model was tested, and the assumptions re-examined. Model fit was measured by the Akaike information criterion[42].

#### Sensitivity Analysis

Sensitivity analysis was performed to measure the impact of assumptions (ii, iii and iv) used to construct the transmission network on the relationship between *ACE2* transcription and number of secondary cases. The assumptions were excluded independently, and the transmission network was iteratively reconstructed with (n-1) assumptions. Additionally, the fourth assumption (iv) was challenged by changing the infectious period from 15 days (three serial intervals) to 10 days (two serial intervals). The number of secondary cases was computed for each transmission network and used in multivariable analysis.

Statistical analysis was performed in R version 4.04 using the packages: readxl, tidyverse, dataexplorer, ggpubr, car, ggsci, stringr, tableone, rio, remotes, lubridate, dplyr, epicontacts, AER, devtools, rlang, DHARMa, MASS, pscl, epiR, EpiCurve[43].

## Results

### Descriptive Statistics

The analytic dataset contains (n = 103) cases of COVID-19 associated with a single-site concurrent series of hospital outbreaks from 07-11-2020 to 04-01-2021 (dd-mm-yyyy) in British Columbia, Canada. Participant characteristics are displayed in Table 1, where the variables of interest are stratified by case description (Patients or Health Care Workers) (Table 1). The bivariate relationship between variables and categorical *ACE2* transcription was also investigated (Table S1). Most cases occurred in patients n = 57 (55%), while n = 48 infections were observed in health care workers (45%). The mean age of patients associated with the hospital outbreaks was 78.61 years, health care workers were significantly younger with a mean age of 41.11 years (P < 0.001). Most of the healthcare workers (n = 41, 89%) and patients were biologic females (n = 33, 58%). *ACE2* transcription and viral load did not differ between case descriptions (P = 0.86, P = 0.30). Six SARS-CoV-2 lineages were characterized by whole genome sequencing, two of which did not have more than one case (B.1.128 and B.1.36.38). The highest proportion of observed SARS-CoV-2 infections were either SARS-CoV-2 lineage AL.1 (n = 56) or B.1.2 (n = 40) (Table 1), these two lineages have at least a five SNP difference between them (Table S4).

**Table 1:** Characteristics of analytic data stratified by case designation (n = 103).

| Variable | Level | Health Care Worker<br>n = 46 | Patient<br>n = 57 | P-Value |
| --- | --- | --- | --- | --- |
| Age (mean (SD)) |  | 41.1 (10.3) | 78.6 (12.1) | <0.001 |
| Biological Sex (%) | Female | 41.0 (89.1) | 33.0 (57.9) | 0.001 |
|  | Male | 5.00 (10.9) | 24.0 (42.1) |  |
| ACE2 Transcription (mean (SD)) |  | 2.11 (2.88) | 2.00 (3.06) | 0.861 |
| RNA Viral Load (mean (SD)) |  | 6.94 (1.79) | 7.33 (1.98) | 0.300 |
| Viral Lineage (%) |  |  |  | 0.385 |
|  | AL.1 | 28.0 (60.9) | 27.0 (47.4) |  |
|  | B.1.128 | 0.00 (0.00) | 1.00 (1.80) |  |
|  | B.1.2 | 14.0 (30.4) | 25 (43.9) |  |
|  | B.1.279 | 0.00 (0.00) | 2.00 (3.50) |  |
|  | B.1.36 | 2.00 (4.30) | 1.00 (1.80) |  |
|  | B.1.36.36 | 1.00 (2.20) | 0.00 (0.00) |  |
|  | B.1.36.38 | 1.00 (2.20) | 0.00 (0.00) |  |

An epidemiological curve was made to show the incidence of positive testers per week over the course of the hospital associate outbreaks. The outbreaks spanned ten epidemiological weeks (Week-#45-2020 to Week-#1-2021) and possesses a non-normal distribution (Figure 1A & B). We are cautious to directly interpret the incidence curve as infection prevention and control practices implemented during the outbreaks (like point prevalence testing and stoppage of admissions) likely biases the observed incidence of SARS-CoV-2 cases overtime. A first peak in cases occurred on Week #46-2020 and a second on Week #52-2020. In the first phase of the outbreaks, many cases occurred on the second floor of the hospital in units 2A, 2B and 2C. Later transmission was predominately observed on floors four or five in units 4B, 4C, 4D and 5A (Figure 1A). Multiple introductions of SARS-CoV-2 into the hospital environment occurred over the surveillance period; however, timely and effective infection prevention and control measures limited transmission of several variants like B.1.36.38, B.1.128 and B.1.279 (Table S3, Figure S2).

**Figure 1:**

Incidence curve of several hospital associated SARS-CoV-2 outbreaks in British Columbia by epidemiological week from 07-11-2020 to 04-01-2021 (n = 103 laboratory confirmed cases). SARS-CoV-2 incidence follows a non-normal distribution where an initial peak caseload was observed in W46-2020 and a second lesser case surge occurred in W52-2020. **A**) SARS-CoV-2 cases are stratified by hospital floor, early in the surveillance period transmission occurred mostly on floor number two and later moved to floors four and five. If the floor where transmission occurred was not determined by the investigating epidemiologist, then it was coded as ‘unknown’ for our analysis. **B)** SARS-CoV-2 cases are stratified by viral variant, n = 7 unique SARS-CoV-2 variants were identified by whole genome sequencing participants diagnostic specimens. SARS-CoV-2 variants AL.1 and B.1.2 caused the highest percentage of cases: AL.1, 55/103, 53% and B.1.2, 39/103, 38%.

### Transmission Network

Permuting all possible transmission pairs in which the primary and secondary case share the same SARS-CoV-2 lineage yielded n = 9389 combinations. Applying the first assumption (i) filtered the possible transmission pairs to n = 2167. The second assumption (ii) eliminated another n = 1480 possible pairs for a total of n = 687. The third and fourth assumptions restricted the possible pairs to n = 382 and n = 76, respectively. A directed transmission network was constructed from n = 103 cases with n = 76 contacts. The network captures n= 55 transmission events for viral lineage AL.1 and n = 39 for B.1.2; transmission of the other viral lineages was either too low (only one case reported) or did not meet the assumption criteria (Figure 2B). The nodes of the transmission network are shown stratified by categorical ACE2 transcription level (Low or High) (Figure 2A). The distribution of *ACE2* transcription among hospital associated cases was tested with a single sample proportion test, 82% of cases had above average *ACE2* transcription (84/103, P<0.001). Transmission pairs were further stratified into primary (n = 34) and secondary cases (n = 28). Primary cases possessed predominately high *ACE2* transcription (29/34, P< 0.001). *ACE2* transcription was similarly enriched in secondary cases (23/28, P<0.01). Overall, 98.6% of transmission events involved at least one case with high *ACE2* transcription (Table S2).

**Figure 2:**

Infection tracing transmission network analysis of a series of hospital associated SARS-CoV-2 outbreaks in British Columbia by epidemiological week from 07-11-2020 to 04-01-2021 (n = 103 laboratory confirmed cases). Nodes represent confirmed cases while the edge show the direction of outgoing transmission, link a primary case with at least one secondary case. **A)** Nodes in the transmission network are stratified by ACE2 transcription (high or low), 82% of cases (84/103) had above average, high ACE2 transcription (P < 0.001). **B)** Nodes in the transmission network are classified by SARS-CoV-2 viral variant. The viral variants AL.1 and B.1.2 caused multiple secondary infections thorough the outbreaks, in the first phase (before Dec.15^th^, 2020) of the study period most cases were AL.1 and later transitioned to B.1.2 in phase two (after Dec.15^th^, 2020).

### Primary Analysis

Multivariable NB regression estimated that a one-unit change in *ACE2* transcription decreases the number of secondary cases in health care workers by −0.13 (95%CI: −0.255 to − 0.0181) adjusting for RNA viral load and case description (Table 2). Effect modification was observed between *ACE2* transcription and case description; in patients, a one-unit change in ACE2 increased the number of secondary cases by 0.187 (95% CI: 0.0101 to 0.370). Therefore, a 5 unit increase in *ACE2* transcription may lead to approximately one more SARS-CoV-2 secondary case per primary patient case. Poisson and quasi-Poisson regression models were constructed using the same variables, neither provided a better fit to the NB option (Table 2).

**Table 2:** Multivariable analysis of the relationship between ACE2 transcription and the number of SARS-CoV-2 secondary cases.

| **Multivariable Regression Models**<br>*(Secondary Cases ~ ACE2 Transcription + RNA Viral Load + Case Description + ACE2 Transcription : Case Description)* |  |  |  |
| --- | --- | --- | --- |
| **Beta Coefficients**<br>*(95% CI)* | **Poisson** | **Quasi-Poisson** | **Negative Binomial**<br>**(NB)** |
| ACE2 Transcription<br>in HCW | -0.164<br>*(-0.236- [-0.0919])* | -0.164<br>*(-0.273-[-0.0528])* | -0.132<br>*(-0.255- [-0.0181])* |
| RNA Viral Load | $2.73 \times 10^{-3}$<br>*($5.34 \times 10^{-3} - 0.0976$)* | $2.73 \times 10^{-3}$<br>*(-0.141-0.150)* | -0.0197<br>*(-0.164- [0.123])* |
| Case (Patient) | -0.808<br>*(-1.20- [-0.421])* | -0.808<br>*(-1.43- [-0.235])* | -0.782<br>*(-1.42- [-0.162])* |
| ACE2 Transcription<br>in Patients | 0.208<br>*(0.0958-0.320)* | 0.208<br>*(0.0382-0.382)* | 0.187<br>*(0.01011-0.370)* |
| AIC | 394 | -- | 346 |
| Theta | 2.44 | 2.44 | 0.922 |
| Score Test<br>(P Value) | P < 0.001 | -- | P = 0.976 |

### Sensitivity Analysis

A sensitivity analysis was performed to measure the impact of assumptions ii, iii and iv on construction of the transmission network and number of secondary cases. The first assumption-that the symptom onset date of the primary case occurs before the secondary case-was not included in the sensitivity analysis, as breaking this assumption would make the secondary case the primary case. Omission of the second assumption (ii), so that the primary case did not share a hospital unit with the secondary case, increased the number of possible transmission pairs to n = 538 (Figure S3). The network was not clearly resolved and no significant relationship between *ACE2* transcription and the number of secondary cases was estimated using a Poisson (B= −0.17 [95%CI: −0.903 to 0.558], P = 0.64) or NB regression model (B = −0.01 [95%CI: −0.082 to 0.0613], P = 0.8) (Figure 3). Both models had a worse fit then when used in the primary analysis (Poisson_AIC_ = 705 and NB_AIC_ = 691). Leaving out the third assumption (iii), to appreciate that the secondary case could have been infected in less time than the average serial interval, increased the number of possible transmission pairs to n = 246 (Figure S4). A significant relationship between *ACE2* transcription and the number of secondary cases was estimated using a Poisson (B = −0.60 [95%CI: −1.08 to −0.125], P = 0.015) and NB regression model (B = −0.11 [95%CI: −0.215 to −0.0189], P = 0.03) (Figure 3). However, neither model fit better then when used in the primary analysis (Poisson_AIC_ = 618 and NB_AIC_ = 551). Re-analysis without the fourth assumption that the infectious period of SARS-CoV-2 does not eclipse 15 days increased the possible transmission pairs to n = 158 (Figure S5), no significant relationship between *ACE2* transcription and the number of secondary cases was estimated using a Poisson (B = 0.0143 [95%CI: −0.284 to 0.313], P = 0.64) or NB regression model (B = 3.90 × 10^−3^ [95%CI: −0.0849 to 0.0911], P = 0.8) (Figure 3). Additionally, assumption four (iv) was challenged by decreasing the infectious period to 9.5 days or ∼ 2 serial intervals (Figure S6, transmission pairs n = 48). Applying a shorter infectious period provided the following estimates using a Poisson (B = −0.270 [95%CI: −0.454 to −0.0852], P = 0.00) and NB regression model (B = −0.160 [95%CI: −0.324 to −0.0118], P = 0.09) (Figure 3). Importantly, the effect modification term was significant in the NB model, and it provided a better fit than when used in the primary analysis (AIC = 269). In patients a one-unit change in ACE2 transcription increases the number of secondary cases by 0.268 (95% CI: 0.0315 to 0.518, P = 0.04), when adjusting for RNA viral load (Figure 3).

**Figure 3:**
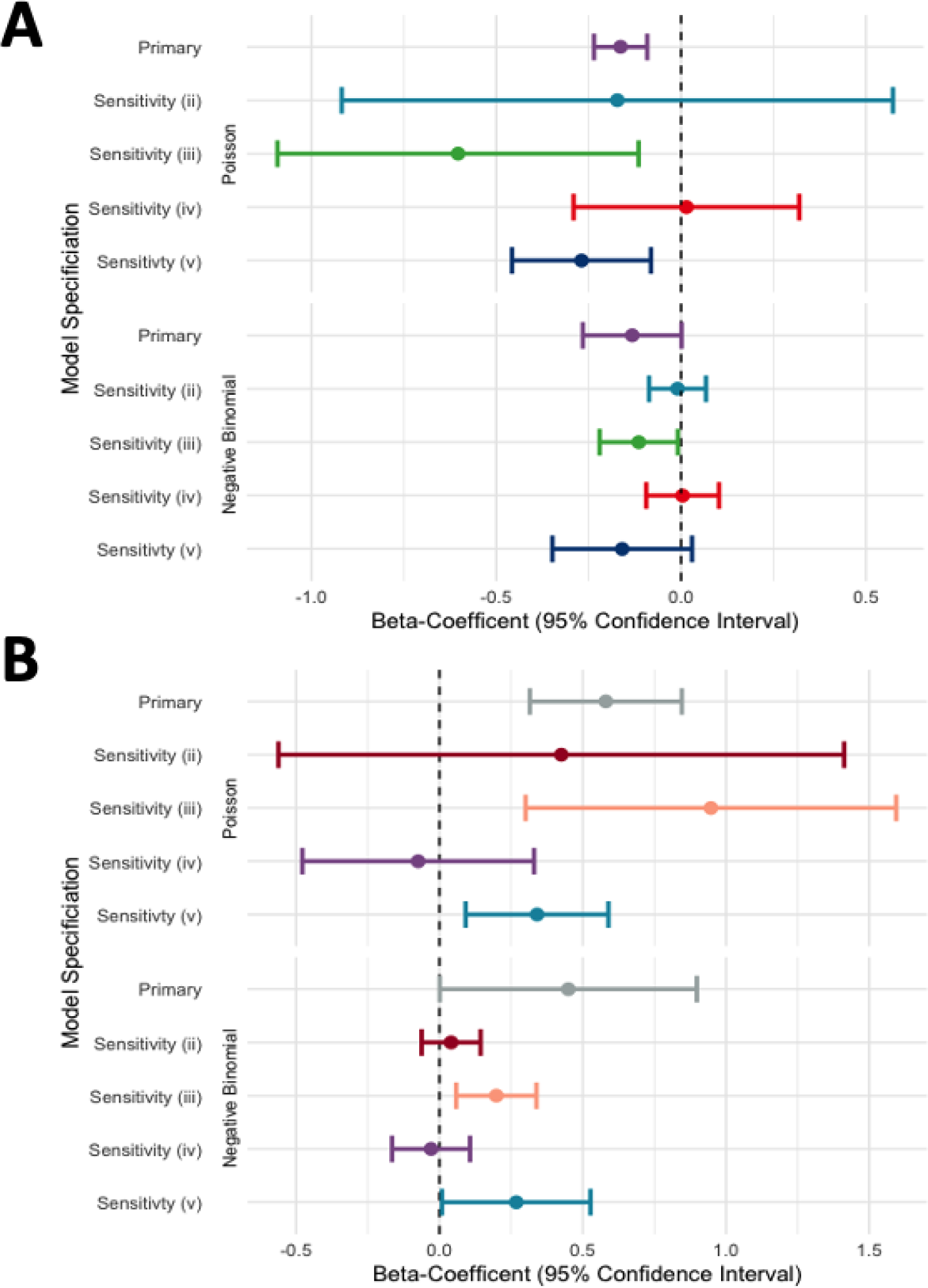
Forest plot of point estimates from the association between nasopharyngeal *ACE2* transcription of the primary case and the SARS-CoV-2 secondary cases. Poisson and Negative Binomial (NB) regression models were used for the primary analysis and sensitivity analysis of the assumptions used to construct the infection tracing networks. **A)** Beta-coefficients and ninety-five percent confidence intervals for the association between nasopharyngeal *ACE2* transcription and secondary cases within health care workers, adjusting for viral load. The best fit NB model shows no association in the primary analysis or sensitivity analysis (ii, iv or v). A significant association between nasopharyngeal *ACE2* transcription and the number of SARS-CoV-2 secondary cases was found for healthcare workers in sensitivity analysis iii, where the serial interval of 5.6 days was not used to build the transmission network. **B)** Beta-coefficients and ninety-five percent confidence intervals for the association between nasopharyngeal *ACE2* transcription and secondary cases within patients, adjusting for viral load. The best fit NB model finds a positive association between in the primary analysis, and sensitivity analysis iii, and v. No association between nasopharyngeal *ACE2* transcription and the number of SARS-CoV-2 secondary cases was found for patients in sensitivity analysis ii and iv, where assumptions of transmission occurring within hospital units and a 15.6-day infectious period were excluded from constructing the transmission network, respectively.

## Discussion

This study describes case-to-case transmission within a series of hospital associated SARS-CoV-2 outbreaks in British Columbia during late 2020 and early 2021. We used a combination of epidemiological data from outbreak reports, laboratory measurements of ACE2, and SARS-CoV-2 lineages derived from whole genome sequencing to recreate the outbreaks using infection tracing transmission network analysis[44]. The transmission network was resolved to provide the possible number of transmission events (secondary cases) per primary case. Bivariate and multivariable analysis was employed to estimate the relationship between nasopharyngeal *ACE2* transcription and the number of potential secondary cases.

Bivariate analysis found that nasopharyngeal *ACE2* transcription did not differ between patients and health care workers. However, *ACE2* transcription was enriched in cases, more than fifty percent of the cases had ‘High’ ACE2. The transmission network constructed with all four assumptions provided n = 76 transmission events for analysis. The simplest recreation of the outbreaks was provided by a sensitivity analysis where the fourth assumption (iv) was changed to stipulate an infectious period of 9.5 days (∼2 serial intervals), which yielded n = 48 transmission events. Multivariable analysis with Poisson regression models was inferior to NB regression as the number of secondary cases was over dispersed. In the primary analysis, NB regression estimated that a one-unit change in nasopharyngeal *ACE2* transcription decreases the number of secondary cases in health care workers by B = −0.132 (95%CI: −0.255 to −0.0181, P = 0.04) and increases transmission in patients by B = 0.187 (95% CI: 0.0101 to 0.370, P = 0.04). Sensitivity analysis using a shorter infectious period of 9.5 days, provided comparable results, no association was found between nasopharyngeal *ACE2* transcription and SARS-CoV-2 transmission from health care workers (P = 0.09). In patients, nasopharyngeal ACE2 expression was associated with secondary transmission B = 0.27 (95% CI: 0.0315 to 0.518, P = 0.04) adjusting for RNA viral load (Figure 3A & B).

Viral transmission exemplifies a complex system where understanding the mechanism of transmission requires deeper analysis than observing the sum of its components. This observational study describes nasopharyngeal *ACE2* transcription as a component of SARS-CoV-2 transmission. In a series of hospital associated outbreaks in British Columbia nasopharyngeal *ACE2* transcription was positively associated with the number of secondary cases (ongoing transmission) in patients but not health care workers. The reason for this difference may stem from health care workers wearing personal protective equipment while providing care. The Public Health Agency of Canada recommends that health care workers providing care to COVID-19 positive patients, as a minimum, adhere to droplet and contact precautions [47][45]. If care requires aerosol generating procedures, then the health care worker dons a N95 respirator with a gown, gloves, and eye protection[45]. Surgical masks and N95 respirators provide a physical barrier between the mucosal membrane of the nasopharyngeal passage and virus carrying droplets or particles suspended in the air. The design and intended use of respirators and surgical masks differ. Respirators are designed to prevent inhalation of airborne particles and must fit tightly to the user’s face. Surgical masks are designed to protect others from aerosolized droplet production from the wearer’s upper respiratory system[46]. In either case, physical barrier provided by a mask can prevent SARS-CoV-2 viral particles from binding to ACE2 proteins on the surface of host cells and initiating the viral replication cycle. Masking may not completely prevent exposure to SARS-CoV-2 yet still decreases the infectious dose below the level necessary for infection[47]. Various studies have demonstrated the utility of surgical masks and N95 respirators at preventing COVID-19 or infection with other respiratory pathogens[47–49]. Use of surgical masks and social distancing was associated with a reduction of 44.9 COVID-19 cases per 1000 students and staff in Boston area school districts. This reduction represents a 4.49% decrease in the incidence of SARS-CoV-2[50]. A clinical trial investigating the use of surgical masks or N95 respirators to prevent SARS-CoV-2 infection in health care workers found no difference in the masks’ efficacy to protect from infection[51]. This suggests that when worn by health care workers, surgical masks and N95 respirators have comparable efficacy to reduce respiratory infections transmitted by small, aerosolized particles <5 μm. Taken together, this evidence supports the hypothesis that health care workers in our study were sufficiently protected by their personal protective equipment to negate the association between nasopharyngeal *ACE2* transcription and secondary transmission. In contrast, hospitalized patients did not wear PPE while receiving care and nasopharyngeal *ACE2* transcription possessed a positive association with secondary transmission. The principle of a mask protecting the wearer from causing transmission instead of protecting contacts from acquiring infection parallels how high nasopharyngeal *ACE2* transcription in an infected person may produce more secondary cases. High nasopharyngeal *ACE2* transcription may promote viral shedding in the upper airway and thus the production of finer aerosols from talking, singing, or sneezing. Fine aerosols can spread further than the large ones, produced lower in the respiratory tract, and pose an increased risk of infection to contacts[52]. As PPE use was not measured in our study competing hypotheses could explain the apparent difference in the relationship between *ACE2* transcription and transmission among health care workers and patients. An unmeasured confounding variable present for health care workers but not patients could bias the relationship between *ACE2* transcription and secondary transmission. For example, health care workers have variable shift lengths and leave the hospital daily after work; therefore, their contact time with an infected patient differs from that of an inpatient who shares the same hospital unit. This example also serves to demonstrate the complexity of measuring SARS-CoV-2 and other respiratory pathogen’s transmission. We acknowledge that SARS-CoV-2 transmission occurs due to an intricate balance of social, biological, and environmental risk-factors of which nasopharyngeal *ACE2* transcription may contribute a singular role[7].

Regulators of *ACE2* transcription in the lower respiratory tract are well documented a lesser evidence base exists for the upper respiratory tract. Importantly, upper, and lower airway ACE2 transcription are not strongly corelated indicating that they may occur independently of each other[53]. In previous work we found that nasopharyngeal transcription of *ACE2* did not differ by age (within adults over the age of eighteen) or biological sex[27], a finding which was further supported by this study. High nasopharyngeal ACE2 expression has been associated with long term inhaled corticosteroid use and exposure to fine particulate matter PM_2.5_. Below average, low nasopharyngeal *ACE2* transcription is associated with type 2 inflammation mediated by the cytokines IL-4, IL-5, and IL-13[54,55]. Therefore, individuals with asthma or allergic rhinitis may have lower upper airway *ACE2* expression than those that do not. Further studies are required to understand additional predictors of upper airway ACE2 expression and if it relates to that of the lower airway.

### Strengths and limitations

The described study has several limitations in design, data collection and analysis. Sampling cases of SARS-CoV-2 but not their contacts prevented us from constructing a more robust transmission network[44]. Contact tracing data would benefit our analysis by increasing power of the study and allow estimation of the relationship between nasopharyngeal *ACE2* transcription and lack of transmission. Estimating the relationship between nasopharyngeal *ACE2* transcription and secondary transmission in only confirmed cases may have resulted in selection bias, where we selected for *ACE2* measurements in persons that had been exposed and infected by SARS-CoV-2[56]. Sampling from a population of hospitalized patients may also have introduced Berkson’s bias (admission rate bias) into the study and restricts our ability to generalize the results to other sub-groups or the community[56]. The hospitalized patients could share an unmeasured exposure, comorbidity or drug treatment which unknowingly affected their nasopharyngeal *ACE2* transcription. Performing a sensitivity analysis helped us understand the importance of our assumptions in building the infection tracing transmission network. The assumption that the primary and secondary case shared the same hospital unit, proved the most important. Without knowing the place of exposure, the transmission network would not have resolved transmission pairs. Whole genome sequencing provided limited specificity of transmission events when the SARS-CoV-2 outbreak was short and genomes were classified by lineage and not at the resolution of single nucleotide polymorphisms (SNPs). Incorporating SNPs into the analysis may increase resolution of the transmission network by more accurately determining transmission pairs. Using RT-qPCR to measure *ACE2* transcription and RNA viral load overapproximates available ACE2 protein and viral particles[23]. Future work should aim to re-estimate the association between nasopharyngeal *ACE2* transcription and secondary cases using data from a SARS-CoV-2 household transmission study. The design of a household transmission study has the benefits of selecting participants from a non-hospitalized population, observing secondary infections and contacts within family clusters in a context with limited use of personal protective equipment.

### Conclusion

We estimate the association between nasopharyngeal *ACE2* transcription and secondary SARS-CoV-2 transmission in a series of hospital associated outbreaks in British Columbia from late 2020 to early 2021. Analysis shows that 98.6% of transmission pairs and 85% of primary cases had high nasopharyngeal *ACE2* transcription. Multivariable analysis adjusting for RNA viral load and interaction by case description found that nasopharyngeal *ACE2* transcription was positively associated with SARS-CoV-2 transmission in hospital patients but not health care workers. We postulate that use of masks among health care workers explains this difference, transmission from health care workers with high ACE2 was interrupted by barrier protection. SARS-CoV-2 transmission remains a complex and unresolved phenomenon driven by biological, environmental, and social risk factors. Differential nasopharyngeal *ACE2* transcription has been observed in COVID-19 negative persons and having high-above average-expression of ACE2 may serve as a risk factor for transmission of SARS-CoV-2.

## Data Availability

The de-identified participant data that support the findings of this study are available from the British Columbia Ministry of Health. Restrictions apply to the availability of these data, which were used under agreement at the British Columbia Center for Disease Control for this study.

## Author Contributions

A.M.N., K.S.K., K.S. and I.S. conceived and designed the study. Epidemiological reporting, data recording and stewardship was performed by K.S. and S.R. Laboratory data reporting and stewardship was performed by N.P. and A.N.J. Laboratory measurements were taken by A.M.N and K.S.K. Data analysis was performed by A.M.N and K.S.K., D.M.P and M.A.I assisted in interpretation of the results. The manuscript was written by A.M.N, all authors helped with editing and provided their approval for publication.

