## Supplementary Data for "Nasopharyngeal Angiotensin Converting Enzyme 2 (ACE2) Expression as a Risk-Factor for SARS-CoV-2 Transmission in Concurrent Hospital Associated Outbreaks"

**Supplementary Materials:**

**Table S1:** Characteristics of analytic data stratified by above ‘High’ or below average ‘Low’ nasopharyngeal *ACE2* transcription (n = 103).

|  | |  | **Low ACE2** | | | **High ACE2** | **P-Value** |
| --- | --- | --- | --- | --- | --- | --- | --- |
| **Variable** | | **Level** | n = 27 | | | n = 76 |  |
| Age (mean (SD)) | |  | 64.89 (20.35) | | | 60.79 (22.42) | 0.406 |
| Biological Sex (%) | | Female | 18 (66.7) | | | 56 (73.7) | 0.655 |
|  | | Male | 9 (33.3) | | | 20 (26.3) |  |
| Case Designation (mean (SD)) | | Health Care Worker | 11 (40.7) | | | 35 (46.1) | 0.801 |
|  | | Patient | 16 (59.3) | | | 41 (53.9) |  |
| RNA Viral Load (mean (SD)) | |  | 6.45 (1.74) | | | 7.40 (1.90) | 0.024 |
| Viral Lineage (%) | | AL.1 | 11 (40.7) | | | 44 (57.9) | 0.011 |
|  | | B.1.128 | 1 (3.7) | | | 0 (0.0) |  |
|  | | B.1.2 | 9 (33.3) | | | 30 (39.5) |  |
|  | | B.1.279 | 1 (3.7) | | | 1 (1.3) |  |
|  | | B.1.36 | 3 (11.1) | | | 0 (0.0) |  |
|  | | B.1.36.36 | 1 (3.7) | | | 1 (1.3) |  |
|  | | B.1.36.38 | 1 (3.7) | | | 0 (0.0) |  |

Variables included in the study are stratified by ‘High’ or ‘Low’ nasopharyngeal *ACE2* transcription. P-values for parametric statistical tests performed on continuous (t-test) or categorical data (X^2^ test) are reported. Viral lineage calls are from whole genome sequencing data classified using the PANGOLIN tool (Version 1.15.1). Data is also available stratified by case designation ‘Health Care Worker’ or ‘Patient’ (Table 1).

**Table S2:** Transmission Network Case Count Contingency Table by ‘High’ or ‘Low’ Nasopharyngeal *ACE2* Transcription.

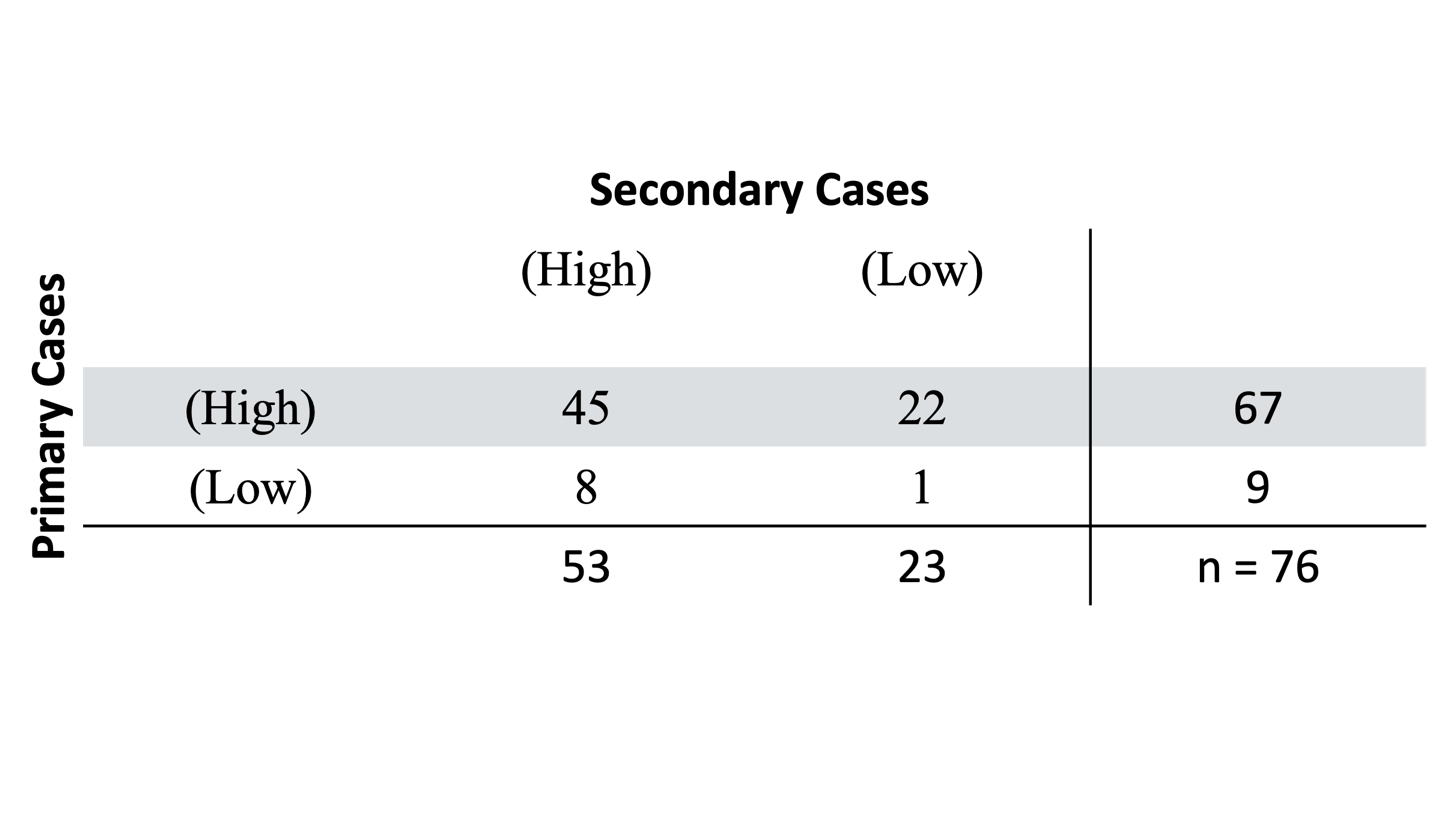

**Table S3:** Descriptive Summary of Single-Site Concurrent Hospital Associated Outbreaks.

| **Outbreak Count** | **Setting (Floor\|Unit)** | **Major Viral Lineage** | **Proportion of Cases with Major Lineage** | **Index Case Collection Date (dd-mm-yyyy)** | **Time Until Last Associated Positive Test (Days)** |
| --- | --- | --- | --- | --- | --- |
| 1 | 2\|A | AL.1 | 18/21 | xx-11-2020 | 10 |
| 2 | 2\|B | B.1.2 | 3/3 | xx-11-2020 | 27 |
| 3 | 2\|B | AL.1 | 6/7 | xx-11-2020 | 16 |
| 4 | 2\|C | AL.1 | 9/17 | xx-11-2020 | 19 |
| 5 | 3\|B | AL.1 | 5/10 | xx-11-2020 | 24 |
| 6 | 3\|C | B.1.2 | 2/2 | xx-11-2020 | 5 |
| 7 | 4\|BA | AL.1 | 2/2 | xx-12-2020 | 6 |
| 8 | 4\|BA | B.1.2 | 4/4 | xx-12-2020 | 18 |
| 9 | 5A | AL.1 | 4/4 | xx-12-2020 | 31 |
| 10 | 5A | B.1.2 | 9/10 | xx-11-2020 | 45 |
| Unassigned Cases | Unknown | AL.1 | 10/18 | xx-11-2020 | 53 |

Included Participants n = 202

Excluded: specimen not identified n = 22

Excluded: sequencing failed n = 63

Excluded: ACE2 transcription was not measured n = 14

Analytic Dataset (Complete Case Data) n = 103

**Figure S1:** Exclusion criteria applied to select an analytic dataset where all included participants had complete case data (no missingness).

**Table S4:** SARS-CoV-2 Viral Variant Whole-Genome Single Nucleotide Polymorphisms.

| **Viral Variant Lineage** | **SNP Count** | **Whole Genome - Unique Single Nucleotide Polymorphisms (SNPs)** | **Closest Relative in the Study** |
| --- | --- | --- | --- |
| AL.1 | 9 | G3716A,C9430T,C9451T,C9561T,C11109T,C13774T,A21625G,G25855T,G28857T | B.1.128 |
| B.1.36 | 12 | C186T,G4180T,G7936T,T10421A,T15747C,C16092T,G18756T,C23415T,G25855T,G26062T,G29254T | B.1.36.36 |
| B.1.128 | 6 | A3735C,A4336G,C5730T,C22264T,T25695C | AL.1 |
| B.1.2 | 7 | G4444T,G17478A,G18315A,C19560T,G23900C,C27942T | B.1.279 |
| B.1.36.38 | 10 | C1572T,G3320A,C9051T,T12101A,G19549T,C20574T,C26395A,C26789A | B.1.36.36 |
| B.1.36.36 | 2 | A13913G,C27911T | B.1.36.38 |
| B.1.279 | 15 | C1909T,C1997T,A3405G,C6255T,C7039T,C11494T,C14177T,G18651T,C24809T,G25234T,A26059G,G28907A,C29733T | B.1.2 |

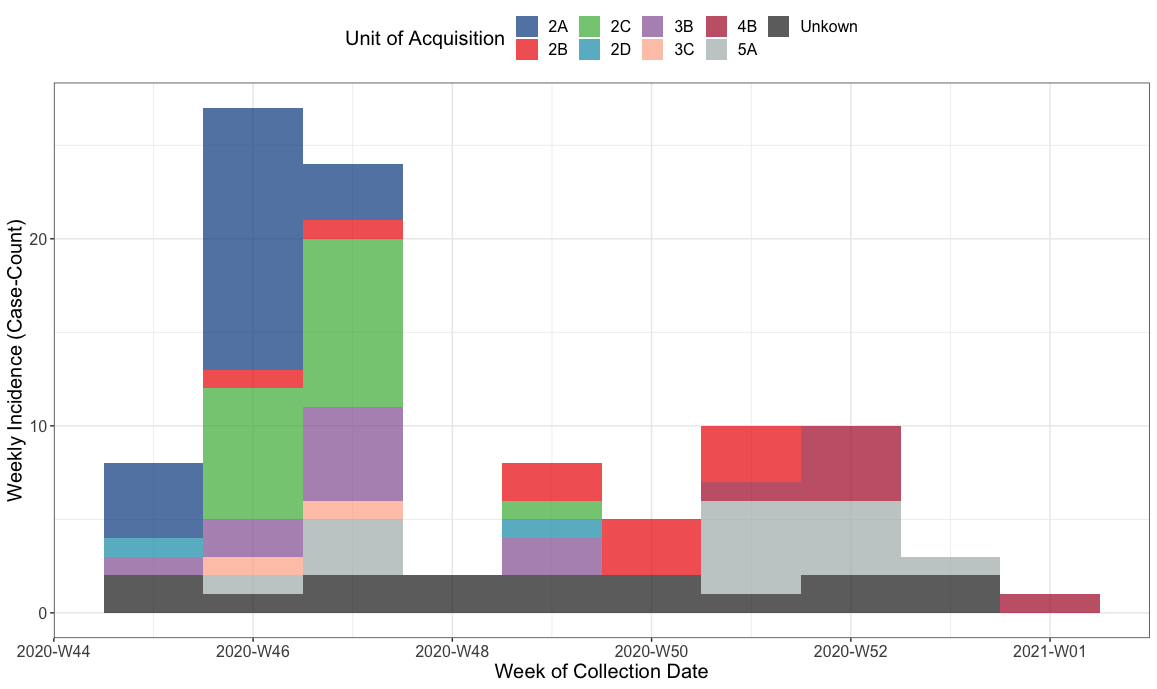

**Figure S2:** Incidence curve for concurrent hospital associated outbreaks stratified by hospital unit (setting) by epidemiological week from 07-11-2020 to 04-01-2021 (n = 103 laboratory confirmed cases). Outbreaks were defined by hospital unit of acquisition, over the surveillance period the hospital declared ~ 10 outbreaks (Table S3), in n = 18, 17% of cases the unit of acquisition was not determined by epidemiological investigation (unkown).

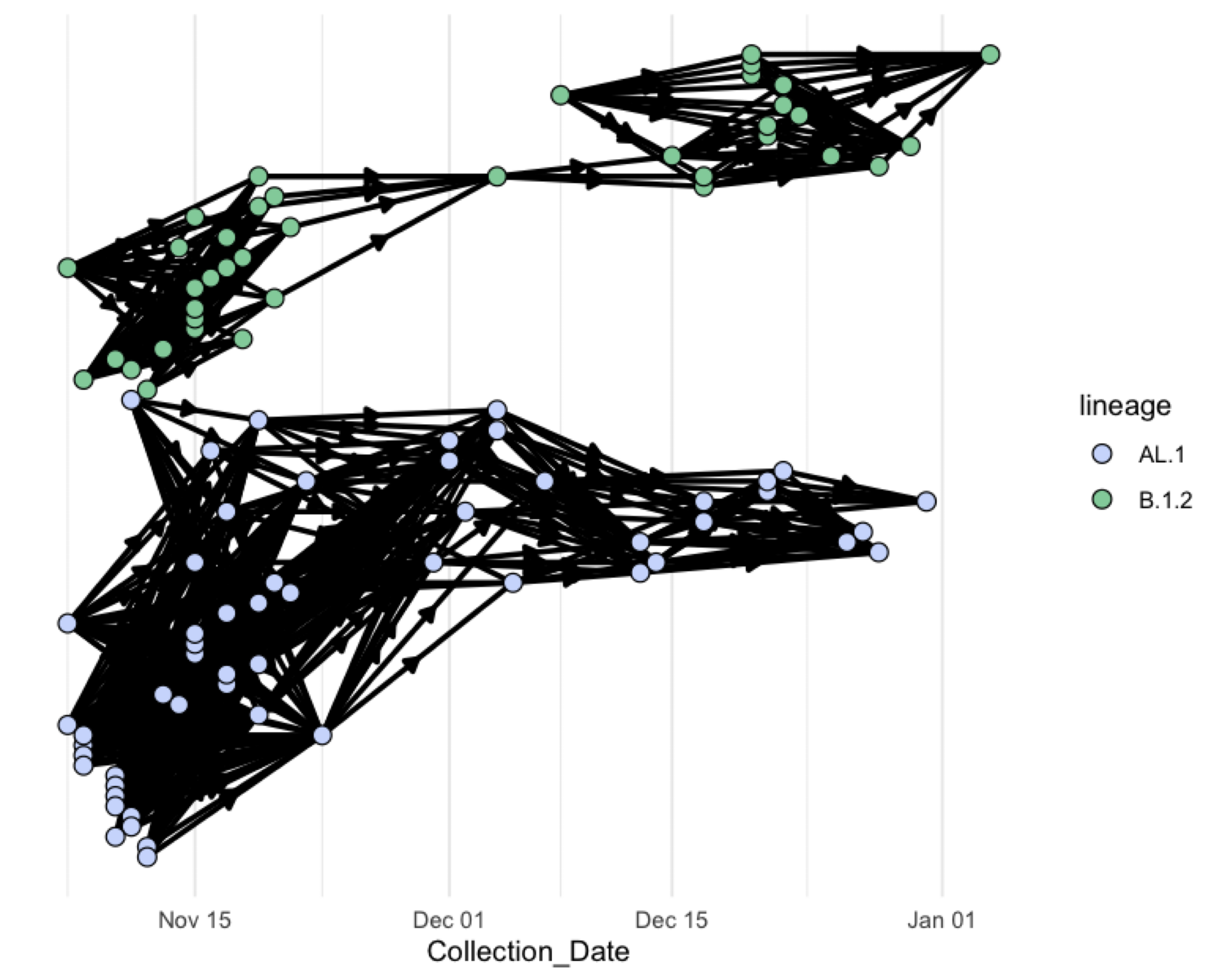

**Figure S3:** Sensitivity analysis of the infection tracing transmission network where the second assumption was omitted, the primary and secondary case do not have to share the same unit of acquisition.

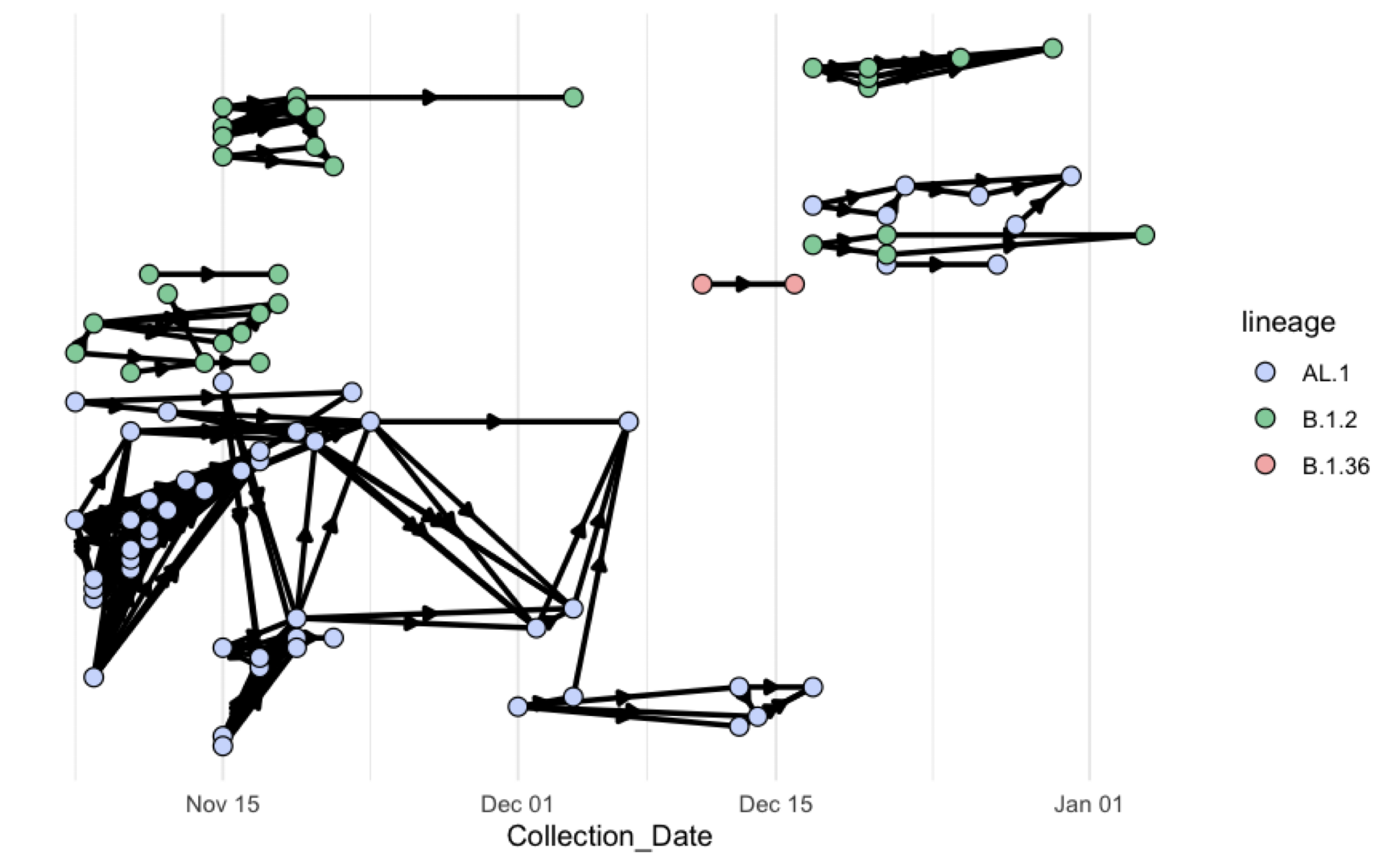
 **Figure S4:** Sensitivity analysis of the infection tracing transmission network where the third assumption was omitted, the collection date of specimens from the primary and secondary case can be less than ~5 days apart.

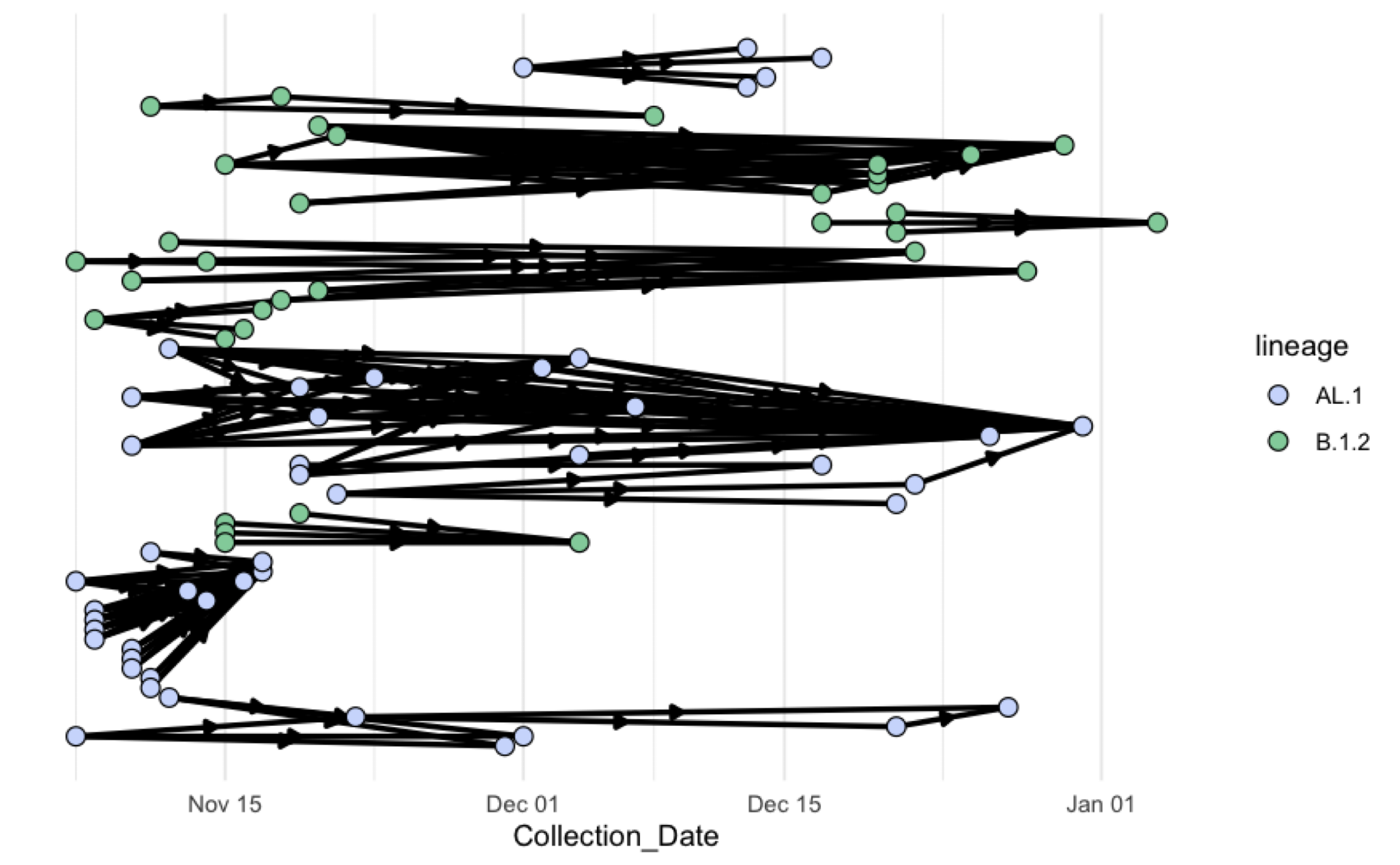

**Figure S5:** Sensitivity analysis of the infection tracing transmission network where the fourth assumption was changed, the collection date of the secondary case is at most three serial intervals (~16 days) from that of the primary case.

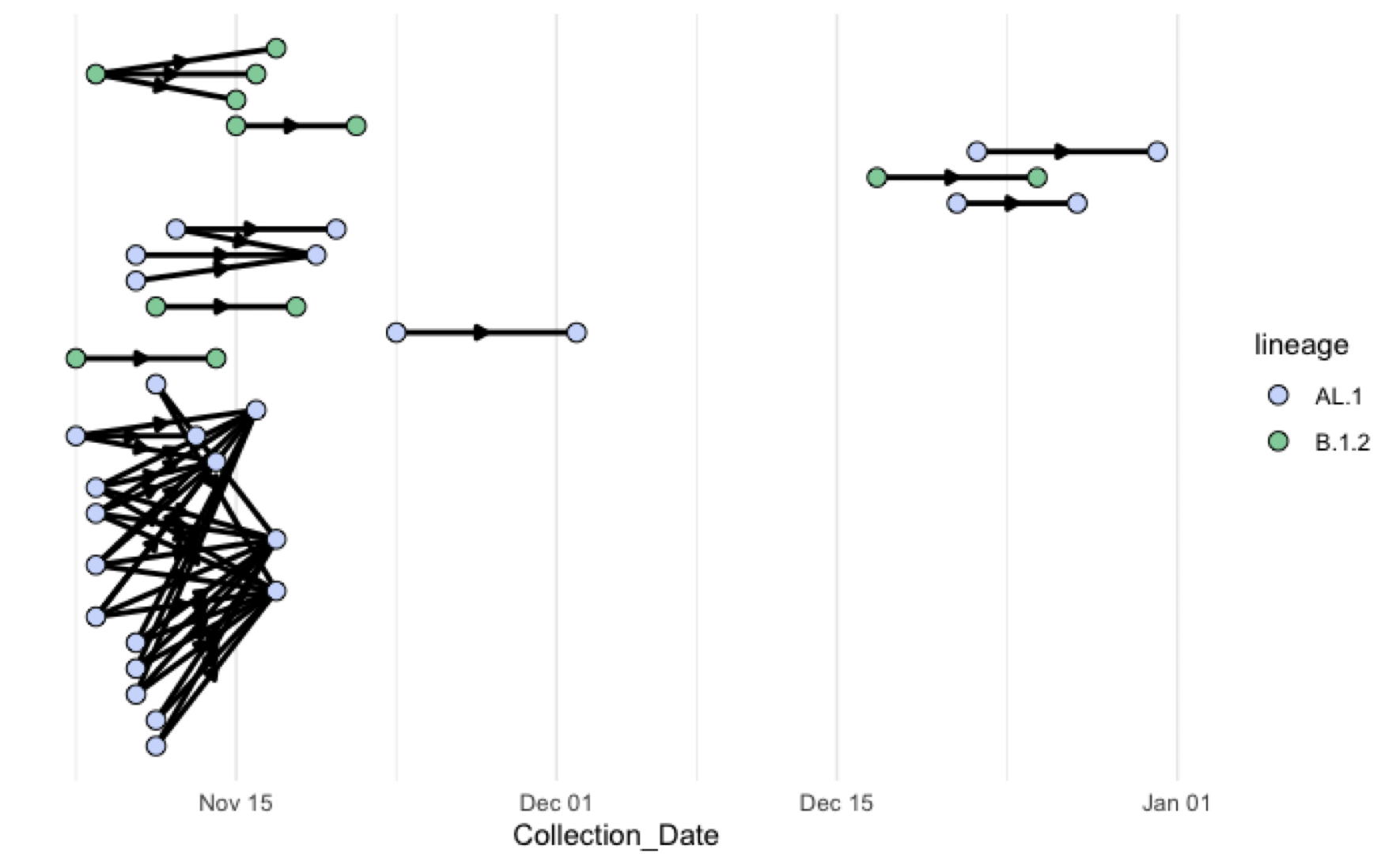

**Figure S6:** Sensitivity analysis of the infection tracing transmission network where the fourth assumption was changed, the collection date of the secondary case is at most two serial intervals (~10 days) from that of the primary case.
